# Trends and prediction in daily incidence of novel coronavirus infection in China, Hubei Province and Wuhan City: an application of Farr’s law

**DOI:** 10.1101/2020.02.19.20025148

**Authors:** Jie Xu, Yajiao Cheng, Xiaoling Yuan, Wei V. Li, Lanjing Zhang

## Abstract

**Background:** The recent outbreak of novel coronavirus (2019-nCoV) has infected tens of thousands of patients in China. Studies have forecasted future trends of the incidence of 2019-nCoV infection, but appeared unsuccessful. Farr’s law is a classic epidemiology theory/practice for predicting epidemics. Therefore, we used and validated a model based on Farr’s law to predict the daily-incidence of 2019-nCoV infection in China and 2 regions of high-incidence.

**Methods:** We extracted the 2019-nCoV incidence data of China, Hubei Province and Wuhan City from websites of the Chinese and Hubei health commissions. A model based on Farr’s law was developed using the data available on Feb. 8, 2020, and used to predict daily-incidence of 2019-nCoV infection in China, Hubei Province and Wuhan City afterward.

**Results:** We observed 50,995 (37001 on or before Feb. 8) incident cases in China from January 16 to February 15, 2020. The daily-incidence has peaked in China, Hubei Providence and Wuhan City, but with different downward slopes. If no major changes occur, our model shows that the daily-incidence of 2019-nCoV will drop to single-digit by February 25 for China and Hubei Province, but by March 8 for Wuhan city. However, predicted 75% confidence intervals of daily-incidence in all 3 regions of interest had an upward trend. The predicted trends overall match the prospectively-collected data, confirming usefulness of these models.

**Conclusions:** This study shows the daily-incidence of 2019-nCoV in China, Hubei Province and Wuhan City has reached the peak and was decreasing. However, there is a possibility of upward trend.

---

The recent outbreak of novel coronavirus (2019-nCoV) in China has infected about 38,800 patients, and claimed 1113 lives. Studies have forecasted future trends of the infection incidence of coronavirus,^1^ but appeared unsuccessful, likely owing to the recently-implemented aggressive interventions.^2^ Farr’s law is a classic epidemiology theory/practice for predicting epidemics.^3,4^ Therefore, we used and validated a model based on Farr’s law to predict the daily-incidence of 2019-nCoV infection in China and 2 regions of high-incidence.

## Methods

We extracted the 2019-nCoV incidence data of China, Hubei Province and Wuhan City from websites of the Chinese and Hubei health commissions, respectively. The model was developed based on the data available on Feb. 8, 2020, and compared with the prospectively collected data afterward. According to prior reports,^4,5^ the ratio 1 was the ratio of a given day’s incidence over that of the day before. Dividing the ratio 1 of one day and the day before resulted in ratio 2. The possible normality of the ratio 2 was examined using Skewness and Kurtosis tests after various data transformations. After identifying the best-fit data transformation format, we examined the potential linear and log-linear associations of incidence with time. Assuming the future ratio 1’s and 2’s would be the same as the mean of the past five dates and ratio 2’s the same as the mean of the last 10 days, we predicted the ratio 1’s and daily incidence. Statistical analyses were conducted using Stata (version 15) and Joinpoint (NCI). All P values were two-sided and a P<.05 was considered statistically significant. The study is exempt from institutional review board’s review due to the use of publically available and de-identified data.

## Results

The daily-incidence was available for China from January 16 to February 11, 2020, for Wuhan and Hubei from January 11 to February 11. Among various data transformation formats, natural-logarithm-transformed incidence data were of normal distribution, without significant log-linear or linear association. The mean of ratio 2 before February 8, 2020 was 0.944 (quartile 0.886-1.051) for China, 0.982 (quartile 0.744-1.317) for Wuhan and 0.948 (quartile 0.768-1.207) for Hubei Province. The future incidence was predicted based on the mean or quartiles of ratio 2 and subsequently inferred ratio 1’s. The daily-incidence significantly decreased after February 8 in all regions, and would continue decreasing until reaching zero on February 25. Given the lower quartile (25%) of ratio 2, the incidence may reach zero by February 20, while based on the upper quartile (75%) of ratio 2, the daily-incidence will keep increasing and reach 3000 on February 25 (**Figure**). Hubei Province and Wuhan City had a similar downward trend (**Table**). The prospectively collected daily-incidence after February 8 appeared to fall in the predicted quartiles in all 3 selected regions.

**Figure.**
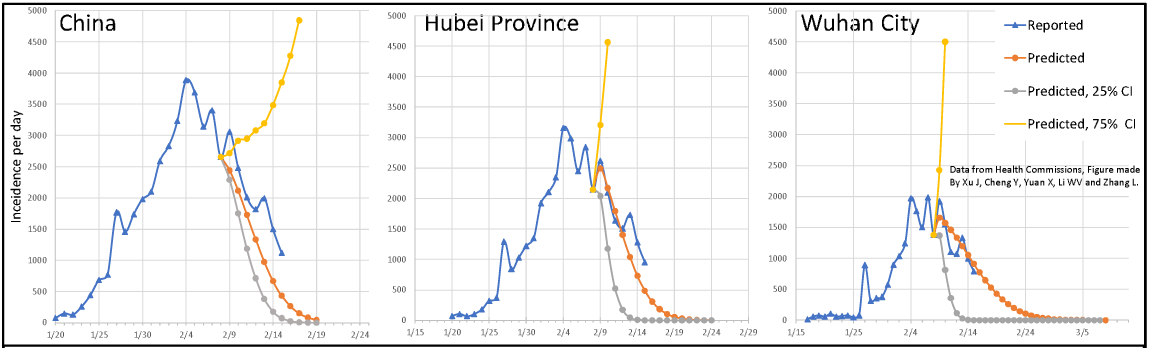
The reported and predicted daily incidence of 2019 novel coronavirus infection in China, Hubei Province and Wuhan City. The predicted daily incidence would reach zero on February 25 for China and Hubei Province, but on March 8 for Wuhan City, due to different ratio 2’s. CI, confidence interval.

**Table.**
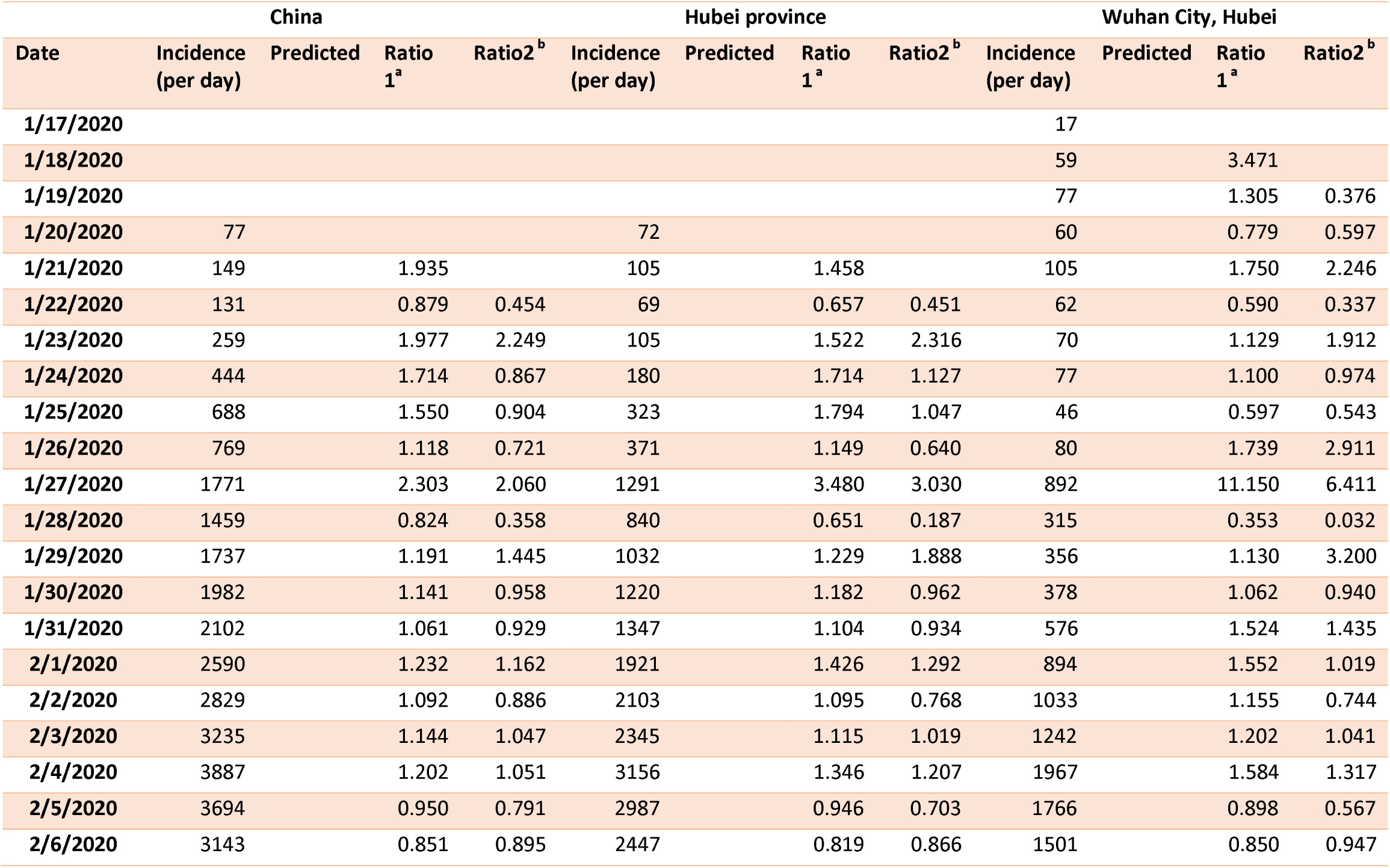
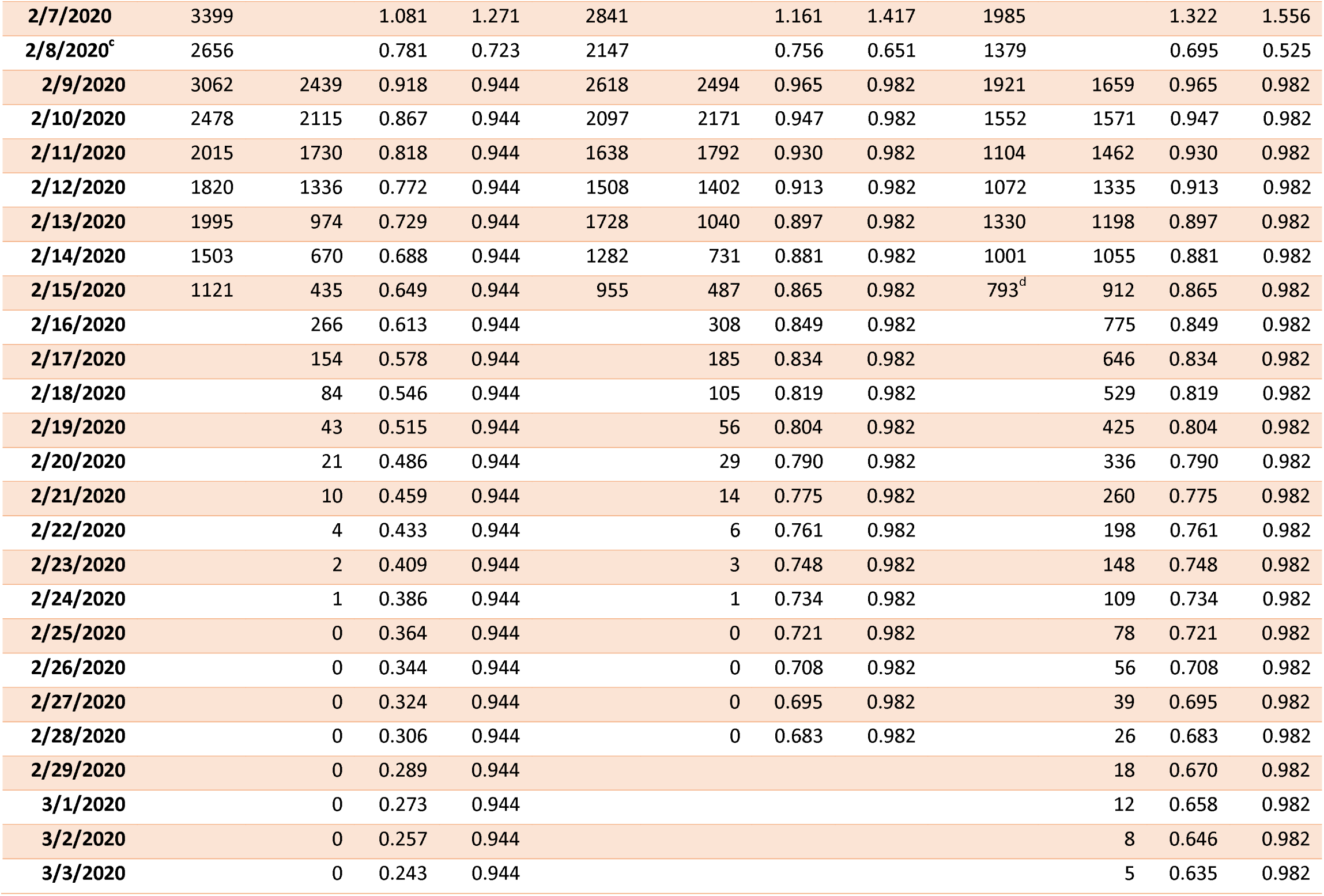
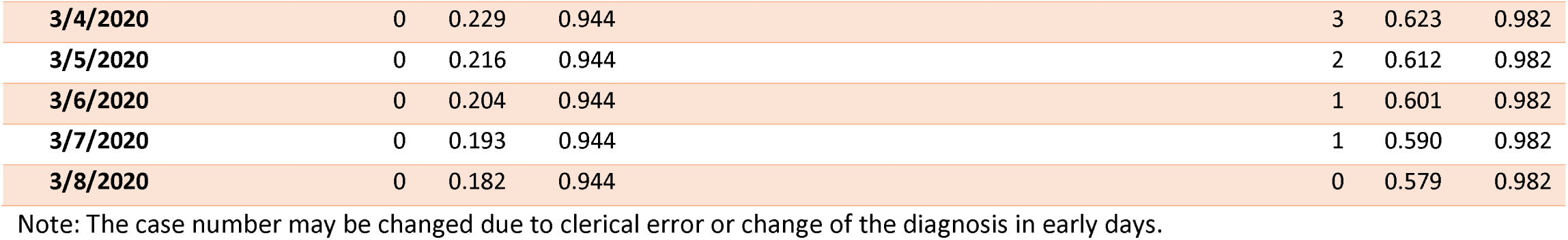
Daily incidence of diagnosed novel coronavirus cases in China and selected areas, Jan. 17 to Feb. 11, 2020, with prediction to March 8, 2020. By Xu J, Cheng Y, Yuan X, Li WV and Zhang L.

## Discussion

Mostly using Farr’s law, we estimated the quartiles of 2019-nCoV daily-incidence in China, Hubei Province and Wuhan City, and predicted the daily-incidence of these regions. Our results imply that the daily-incidence has reached its peak and will likely decrease continuously in China, Hubei Province and Wuhan City, while the predicted 75% CI of their daily-incidence had an upward trend. Therefore, governments, healthcare providers and residents should be cautiously optimistic, and recognize the potential upward trend of daily-incidence. A rapid growth in daily-incidence may occur when the residents of high-prevalence regions, such as Hubei Province, return to their workplaces in low-prevalence regions at the end of Chinese New-Year holidays. Extreme caution thus should be used to prevent such an upward trend.

Limitation includes possible oversimplification of the disease’s natural history by this model. However, our prospectively-collected data and recent works proved the usefulness of the Farr’s law.^5^ This simple yet powerful method has also successfully predicted the trends in incidence of opioid overdose in the U.S.^4^

## Data Availability

All data and their updates are available at https://github.com/thezhanglab/coronavirus

## Acknowledgement

We salute and thank our teachers, classmates and colleagues, fellow residents and public servants, who fiercely and tirelessly fought against the novel coronavirus outbreak in Wuhan and other parts of China. The data will be regularly updated at https://github.com/thezhanglab/coronavirus, as new prospectively collected incidence data become available.

## Author Contributions

Drs Xu and Zhang had full access to all of the data in the study and equally takes responsibility for the integrity of the data and the accuracy of the data analysis. They are both senior authors. Drs Xu and Cheng both contributed equally and should be considered co-first authors.

Concept and design: Xu, Zhang.

Acquisition, analysis, or interpretation of data: All authors.

Drafting of the manuscript: Xu.

Critical revision of the manuscript for important intellectual content: All authors.

Statistical analysis: Li, Zhang.

Supervision: Xu, Zhang.

## Bibliography

1. Wu JT, Leung K, Leung GM. Nowcasting and forecasting the potential domestic and international spread of the 2019- nCoV outbreak originating in Wuhan, China: a modelling study. Lancet. 2020 doi:10.1016/S0140-6736(20)30260-9.

2. News B. Coronavirus: Wuhan shuts public transport over outbreak. 2020; https://web.archive.org/save/ https://www.bbc.com/news/world-asia-5.china-51215348. Accessed Feb. 9, 2020.

3. Bregman DJ, Langmuir AD. Farr’s law applied to AIDS projections. JAMA. 1990;263(11):1522–1525.

4. Darakjy S, Brady JE, DiMaggio CJ, Li G. Applying Farr’s Law to project the drug overdose mortality epidemic in the United States. Inj Epidemiol. 2014;1(1):31. doi:10.1186/s40621-014-0031-2.

5. Santillana M, Tuite A, Nasserie T, et al. Relatedness of the incidence decay with exponential adjustment (IDEA) model, “Farr’s law” and SIR. compartmental difference equation models. Infect Dis Model. 2018;3:1-.12. doi:10.1016/j.idm.2018.03.001.

